# OxCOVID19 Database, a multimodal data repository for better understanding the global impact of COVID-19

**DOI:** 10.1101/2020.08.18.20177147

**Authors:** Adam Mahdi, Piotr Błaszczyk, Paweł Dłotko, Dario Salvi, Tak-Shing Chan, John Harvey, Davide Gurnari, Yue Wu, Ahmad Farhat, Niklas Hellmer, Alexander Zarebski, Bernie Hogan, Lionel Tarassenko

## Abstract

Oxford COVID-19 Database (OxCOVID19 Database) is a comprehensive source of information related to the COVID-19 pandemic. This relational database contains time-series data on epidemiology, government responses, mobility, weather and more across time and space for all countries at the national level, and for more than 50 countries at the regional level. It is curated from a variety of (wherever available) official sources. Its purpose is to facilitate the analysis of the spread of SARS-CoV-2 virus and to assess the effects of non-pharmaceutical interventions to reduce the impact of the pandemic. Our database is a freely available, daily updated tool that provides unified and granular information across geographical regions.

## Background & Summary

Characterising the impact of the COVID-19 pandemic and understanding the efficacy of policy interventions requires a comprehensive, well-formatted and easily accessible database. The World Health Organisation and the European Centre for Disease Prevention and Control collect daily statistics about cases and deaths from governmental sources. These aggregated databases are widely used in research but they lack granularity and context. In addition, several academic institutions have curated high quality data sets aiming at capturing variables not included in the aforementioned databases: the Coronavirus Resource Center at John Hopkins University^1^, the Real-time Case Tracker^2^, *The Economist*’s Tracker for COVID-19 Excess Deaths^3^and the Oxford COVID-19 Government Response Tracker^4^. In constrained research contexts related to the pandemic, these databases prove to be immensely useful to researchers and policy-makers seeking to understand both the causes of the spread and the efficacy of public health interventions. Linking such heterogeneous data is vital to understanding the context which gave rise to the observations and to making inferences at a finer spatial resolution. However, the process of linking relevant data across these sources is complex and requires great care.

The OxCOVID19 Database aims to link different modalities of data, reported at the national and regional level, including epidemiological information on COVID-19 (confirmed, deaths, recovered, hospitalised, etc.), government response (school closing, economic measures, etc.), mobility (e.g., change in mobility trends of humans in various places), weather (e.g., temperature, humidity, precipitation, etc.), socioeconomic statistics and value surveys (Figure 1). The database uses an established spatial index GID^5^, which spans several administrative levels. Wherever possible, the OxCOVID19 Database draws upon official government sources, work by university-based or government research groups and data from peer-reviewed scientific papers. The data are provided with the different granular spatial level thereby facilitating a better understanding of how regional characteristics inform the spread of the disease. Well-linked and granular data of this type can enable the construction of more accurate models of the pandemic by allowing reliable estimation of the required parameters for relatively small regions, avoiding the process of averaging them on a country level. They also increase our understanding of the efficacy of various interventions at the state and regional levels. Thus, we hope this resource in combination with mathematical modelling and machine learning for data analytics will enhance our understanding of the COVID-19 pandemic and facilitate the development of strategies to reduce the impact on society. Some of the key questions which the OxCOVID19 Database can help to answer include the assessment of the effectiveness of different types of non-pharmaceutical interventions such as government lockdowns, mobility restrictions, and social distancing in reducing the spread of infections^6^.

**Figure 1.**
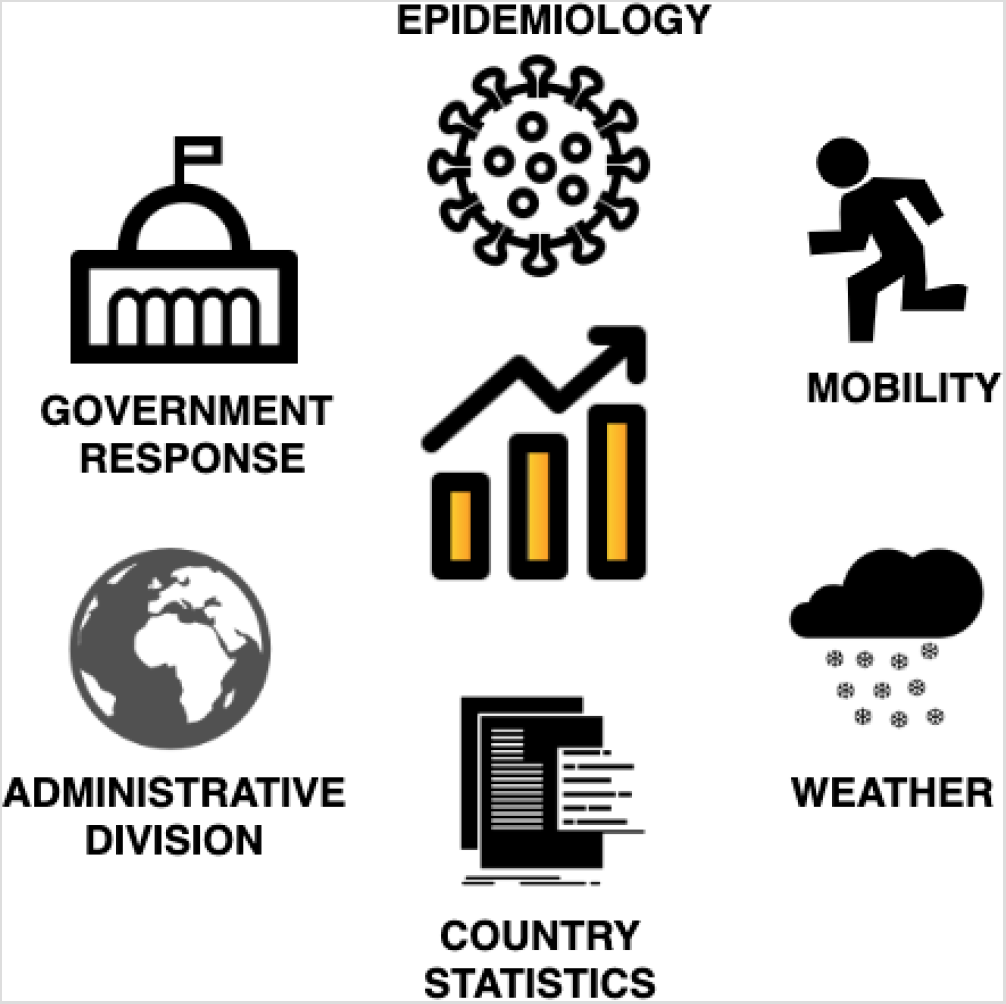
The main types of data categories included in the OxCOVID19 Database.

## Methods

### Data Sources

All the data available in the OxCOVID19 Database are collected from publicly available sources including scientific reports^7^, government press releases, briefings, and similar. For the epidemiological data we relied mostly on official government sources including websites and repositories from Ministries of Health, regional Public Health Authorities, university research groups and official social media accounts. Government response data comes from the Coronavirus Response Tracker as-sembled by researchers from the Blavatnik School of Government at the University of Oxford^4^. For mobility data, we used Community Mobility Reports by Google^8^ and Mobility Trends Reports by Apple^9^. The meteorological data (available from January 1, 2020 onwards) have been made available by the UK Met Office Global Weather Data for COVID-19 Analysis^10^. The socioeconomic statistics and demographics data come from various sources generously made available by the World Values Survey^11^, European Values Study^12^, and the World Bank^13^. A full and updated list of data sources is maintained on https://github.com/covid19db/data.

### Unifying the data across geographical regions

The OxCOVID19 Database links multimodal data for different levels of administrative division. The largest administrative subdivision of a country will be called the “first-level administrative division”, “first administrative level”, or “Level-1” (e.g., “states” in the USA or “voivodeships” in Poland). The next smaller regions will be described as the “second-level administrative division”, “second administrative level” or “Level-2” (e.g., “counties” in the USA, “powiaty” in Poland); similarly “third-level administrative division”, “third administrative level” or “Level-3” (e.g., “gminy” in Poland). Not every country has a third level (e.g., the USA), some countries do not even have a second level, but we include these where available.

We link data from multiple sources and various levels of administrative subdivision into one relational database using the GID from the Global Administrative Areas (GADM) database as a geographical identifier^5^. The goal of the GADM database is to “map the administrative areas of all countries, at all levels of sub-division”. It is freely available for non-commercial use as is the case here. The GID identifies a geographic area with an alphanumeric string. For example, the string ’CHN.16.4_1’ can be decoded as follows: the first three letters are the ISO 3166–1 alpha-3 country code for Mainland China; the ’16’ indicates the Level-1 subdivision, here the province of Jiangxi; the ’4’ represents the Level-2 division, here Jingdezhen City; finally, the ’_1’ indicates a version number for the GID, which only changes in the event of major internal reorganisation and allows for backwards compatibility. To each GID a polygon is associated giving the boundaries of the geographical area to an extremely fine resolution. A dedicated table named ADMINISTRATIVE_DIVISIONS contains the GIDs together with their place names and locations, expressed both as a single point and as a polygon. The resolution of the polygon is reduced from that given in GADM in order to conserve space.

Every record, insofar as possible, is matched to a GID or a list of GIDs. This allows the user to match different modalities of data together using GIDs, including across hierarchies. While GID could potentially be used as the main geographical key in our database, we have chosen to introduce some redundancy and use names (as standardised in GADM^5^) of the regions along with their GIDs for ease of use as well as to permit the exceptional absence of GIDs.

Typically, assigning a GID to the region referred to in a record is a straightforward matter. Slight inconsistencies with spelling variations, prefixes, and suffixes can sometimes be an obstacle to carrying out a direct text match, but this requires only limited and obvious manual adjustment. Some records lack the geographical specificity needed to assign a GID, such as where the administrative subdivision is listed as “Unknown”. There are, however, more challenging situations, often relating to administrative reorganisations which have taken place since the release of GADM 3.6. These are best demonstrated by means of example.

#### Handling of boundary changes – some examples

(i) In UK-England, some local authorities (Level-2 units) have undergone boundary changes, with Level-3 units being moved between or promoted to Level-2 units. Each local authority is therefore easily expressed as a list of Level-2 or Level-3 units.
(ii) A similar situation has occurred in Colombia: The Level-1 Department of Cundinamarca (COL.14_1) contains the Capital District, Bogotá, known in GADM as Santafé de Bogotá (COL.14.79_1). However, Bogotá in fact has the status of an independent department. We can still express both Bogotá and Cundinamarca excluding Bogotá by lists of Level-2 GIDs. Unfortunately, Cundinamarca excluding Bogotá contains 114 Level-2 regions. But, we still express it accurately since any alternative would involve a geographical overlap in reporting and possible ambiguity.
(iii) Norway: A reorganisation of counties (Level-1 units) in Norway resulted in several mergers. However, there were also some minor boundary changes. It is possible, as with Colombia, to express all units as lists of Level-2 GIDs. However, it would be extremely cumbersome, and unlike the case of Colombia it is not necessary to avoid geographical overlap, and would result in little gain in accuracy. For simplicity, lists of Level-1 units are used.

#### New organisational schemes – some examples

Where there has been wholesale reorganisation or reporting takes place according to an organisational schema which is not composed of administrative divisions, the situation is less easily handled.

(iv) UK-Scotland: The reporting regions in Scotland are local health boards, which are not compatible with the administrative units, which are local authorities with Level-2 GIDs. We have endeavoured to represent the health boards as accurately as possible with GIDs.
(v) Latvia: The reorganisation of regions in Latvia resulted in much smaller regions which are not included in GADM. The cities and municipalities of Latvia cannot be associated to GIDs.

### Epidemiology

While our goal is to collect epidemiological data on the regional level for as many countries as possible, we initially sought to prioritise countries to include. To determine priority levels we incorporated three criteria: total population, air traffic volume, and number of COVID-19 related deaths. All countries were ordered according to each of the three criteria on 5 May 2020 and the ranks of countries with respect to each criterion are added to give the priority score (ie, we used a Borda count).

The top 20 countries according to this rank at the time of prioritisation were: United States, China, India, Brazil, United Kingdom, Indonesia, Germany, Turkey, Japan, Spain, Ireland, Russian Federation, France, Italy, Mexico, Pakistan, Belgium, Canada, Iran, Nigeria. We have successfully included regional data for all but Turkey and Iran in the database. At the time of writing, 41 countries have been included at level-1, of which 6 countries are present at level-2, with the United Kingdom at level-3.

### Aggregation of weather variables

The WEATHER table is composed of 47 meteorological variables obtained from the UK Met Office^10^. The variables provide information about temperature, sunshine, humidity and precipitation. This information is sampled daily and reported on a 12 km *×* 12 km uniform latitude-longitude grid.

To provide these data in a manner which permits linking with the other tables, we report the mean value for each variable across all grid points contained in Level-1 and Level-2 GADM subdivisions, along with the standard deviation and the number of grid points in the region. We report on a daily basis starting from 1 January 2020.

This level of subdivision was chosen on the basis that almost all Level-2 regions contain a grid point. Where the region contains no grid point, no record is created. This, however, happens in fewer than 0.5% of the cases. Using Level-3 instead of Level-2 would result in a large number of missing records, while using Level-1 would be overly coarse leading to high standard deviations and reduced explanatory power. Further, values for larger geographical units can be obtained by the user by averaging over the smaller subdivisions taking into account the number of points in each region.

#### World Bank data

The World Bank Development Indicators dataset^13^ are an easily accessible set of country-level indicators including economic characteristics (like GDP), quality of healthcare and other metrics. Each record includes an alpha-3 country code (equivalent to GID) allowing them to be linked to our database. Naturally, not all time series are complete for all countries. For ease of use we provide the latest reported values for all indicators. For the full list of available indicators see https://data.worldbank.org/indicator/.

#### Value surveys

We have extracted a number of indicators from the World Values Survey^11^ and European Values Study^12^ including information on the values and beliefs of people; their trust in government, healthcare and scientific institutions; level of poverty; and similar socioeconomic, political, and demographic indicators.

The statistics are aggregated and equipped with the appropriate GID both at the country level and at a regional level where possible. These regions are generally larger than GADM Level-1 and included only for the same 20 countries which were prioritised for epidemiological data.

Our Integrated Values Surveys dataset is obtained by merging together all fully released waves of the World Values Survey and the European Values Study. There is no official release of this integrated dataset – we merged it following the official guidelines^14^making appropriate adjustments where the guidance has not provided the correct matching.

For each survey question, we report the frequency of each answer. Because each possible answer generates a column, the resulting table has more than 15,000 columns. To reduce the size of the table we instead stored all the statistics for each country/region in a nested dictionary, placed in the column “properties” in the SURVEYS table.

## Data Records

**Figure 2.**
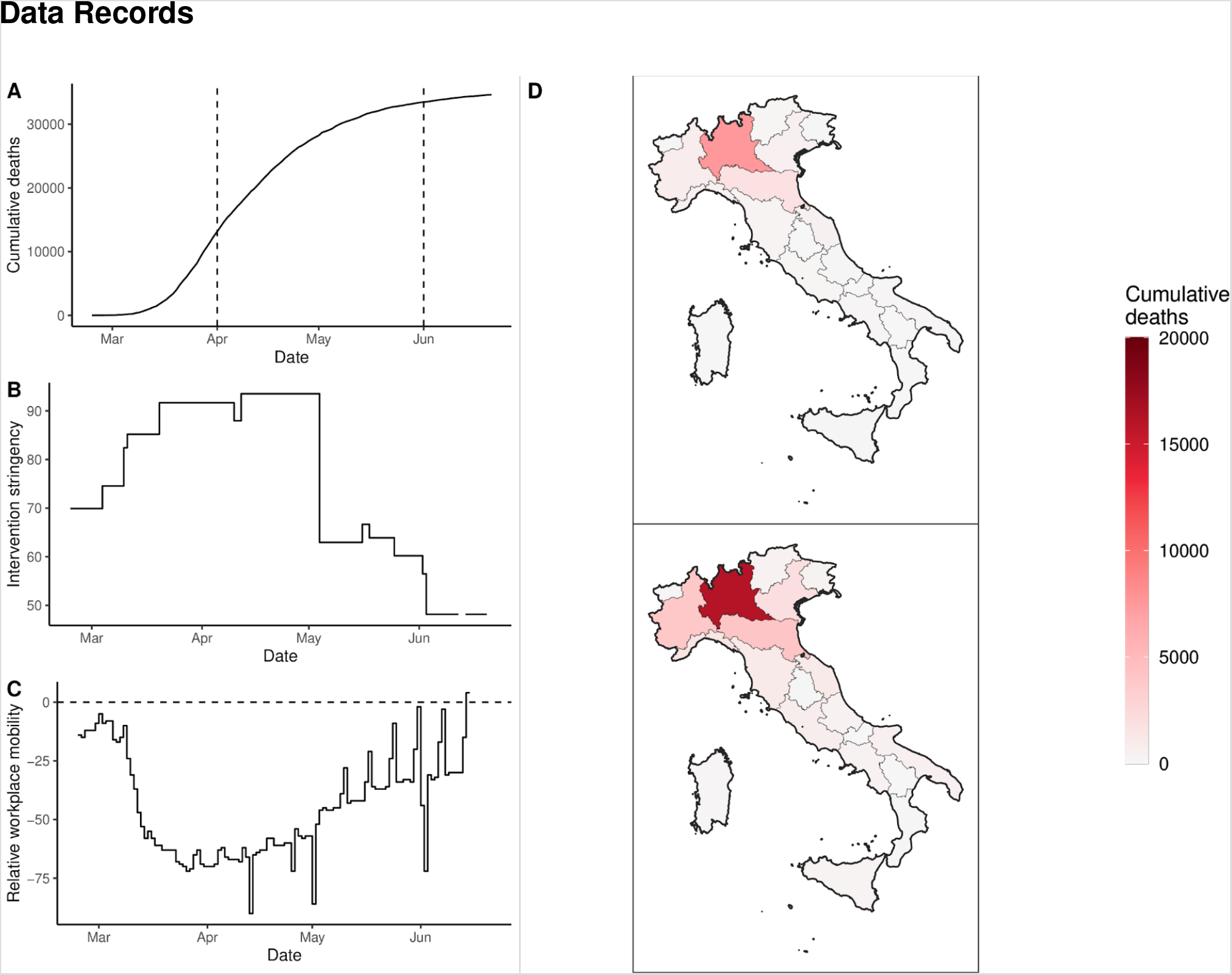
Sample data for Italy demonstrating the data types that are provided by OxCOVID19 Database. Panel A: the cumulative number of deaths through time with two time points (corresponding to 1st of April and 1st of June) indicated by dashed lines. Panel B: the intervention stringency, which is further stratified by the precise type of non-pharmaceutical interventions (see [Hal2020]). Panel C: the relative mobility for workplace activity from Google, the dashed line corresponds to parity with historical values. Panel D: the spatial distribution of the cumulative number of deaths across Italy on the April 1 and June 1, 2020, which corresponds to the dashed lines in Panel A.

The database is available to download at https://covid19.eng.ox.ac.uk/. The data are stored in a PostgreSQL database. CSV extracts from this database are available to access at https://github.com/covid19db/data. A complete archive copy of the database in CSV format as of 31–07–2020 has been stored under the https://doi.org/10.6084/m9.figshare.12746150.

### Common columns for joining tables

The following columns are used to uniquely identify each record and query the database in order to combine different modalities of information: (K1) source – an abbreviation indicating the data source; (K2) **date** – ISO 8601 date (YYYY-MM-DD) of the record under consideration; (K3) **GID**; (K4) **country** – English name for a country as it appears in the GADM database, (K5) **countrycode** – ISO 3166–1 alpha-3 country codes, (K6) **adm_area_1** – specifying first-level administrative country subdivision, (K7) **adm_area_2** – specifying second-level administrative country subdivision, (K8) **adm_area_3** – specifying third-level administrative country subdivision. Note that although (K1)-(K3) uniquely identifies each record the additional columns such as country, countrycode or different levels of administrative division permit more user friendly means to query groups or aggregates of the data as necessary. (K6)-(K8) are strings in the Latin alphabet given as they appear in the GADM database unless the region is not associated with a single GID. In most such cases, it is reported as in the original source. In the case of an Upper Tier Local Authority of England, such as the Boroughs of Greater London, for the sake of ease of use, divisions are listed with their names under **adm_area_2**, with **adm_area_3** being NULL.

### Administrative Divisions

The ADMINISTRATIVE_DIVISIONS table (see Table 1) contains the geographic features and information associated with each GID, extracted from GADM^5^. It includes six linking columns (K3)-(K8), followed by **countrycode_alpha2**, the ISO 3166–1 alpha-2 country code, **adm_level**, specifying which level of division it is, **adm_area_1_code, adm_area_2_code** and **adm_area_3_code**, providing the GID for each higher level administrative division, **properties**, which includes alternative names and identification codes and three geometric features: **latitude** and **longitude**, specifying the centroid of the region, and geometry, specifying the simplified boundaries of the region (shapefiles) for mapping purposes.

**Table 1.** Schema for ADMINISTRATIVE_DIVISIONS table.

| Name | Type | Description |
| --- | --- | --- |
| **gid** | array | Unique geographical ID, for more details see gadm.org |
| **country** | varchar | English name for the country |
| **countrycode** | varchar | ISO 3166-1 alpha-3 country codes |
| **countrycode_alpha2** | varchar | ISO 3166-1 alpha-2 country codes |
| **adm_area_1** | varchar | Level-1 administrative country subdivision |
| **adm_area_1_code** | varchar | First-level administrative country code subdivision |
| **adm_area_2** | varchar | Level-2 administrative country subdivision |
| **adm_area_2_code** | varchar | Second-level administrative country code subdivision |
| **adm_area_3** | varchar | Level-3 administrative country subdivision |
| **adm_area_3_code** | varchar | Third-level administrative country code subdivision |
| **adm_level** | integer | 0 - for countries level, 1 - for regions etc. |
| **latitude** | float | Geographic coordinate of region's centroid |
| **longitude** | float | Geographic coordinate of region's centroid |
| **properties** | json | Additional attributes describing region |
| **geometry** | geometry | Polygon describing geographical area |

### Epidemiological data

The EPIDEMIOLOGY table includes all eight linking columns, (K1)-(K8), followed by **tested** – number of tests; **confirmed** –number of confirmed cases; **dead** – number of deaths; **recovered** – number of individuals recovered; **hospitalised** – number of individuals hospitalised; **hospitalised_icu** – number of individuals in Intensive Care Units; **quarantined** – number of individuals quarantined (see Table 2).

**Table 2.** Schema for EPIDEMIOLOGY table.

| Name | Type | Description |
| --- | --- | --- |
| **source** | varchar | Specify data source |
| **date** | date | Day of the statistics |
| **gid** | array | Unique geographical ID, for more details see gadm.org |
| **country** | varchar | English name for the country |
| **countrycode** | varchar | ISO 3166-1 alpha-3 country codes |
| **adm_area_1** | varchar | Level-1 administrative country subdivision |
| **adm_area_2** | varchar | Level-2 administrative country subdivision |
| **adm_area_3** | varchar | Level-3 administrative country subdivision |
| **tested** | int | Number of people tested |
| **confirmed** | int | Number of confirmed cases |
| **dead** | int | Number of deaths |
| **recovered** | int | Number of confirmed who recovered |
| **hospitalised** | int | Number of confirmed who are/have been hospitalised |
| **hospitalised_icu** | int | Number of confirmed who are/have been in the intensive care |
| **quarantined** | int | Number of confirmed with home quarantine |

### Government response data

The GOVERNMENT_RESPONSE table includes all eight linking columns, (K1)-(K8), followed by a number of indicators (see Table 3), prepared and curated by researchers from the Blavatnik School of Government, University of Oxford^4^. These indicators are grouped into the following categories: containment and closure, economic response and health systems, and miscellaneous policy announcements that do not fit anywhere else.

**Table 3.**
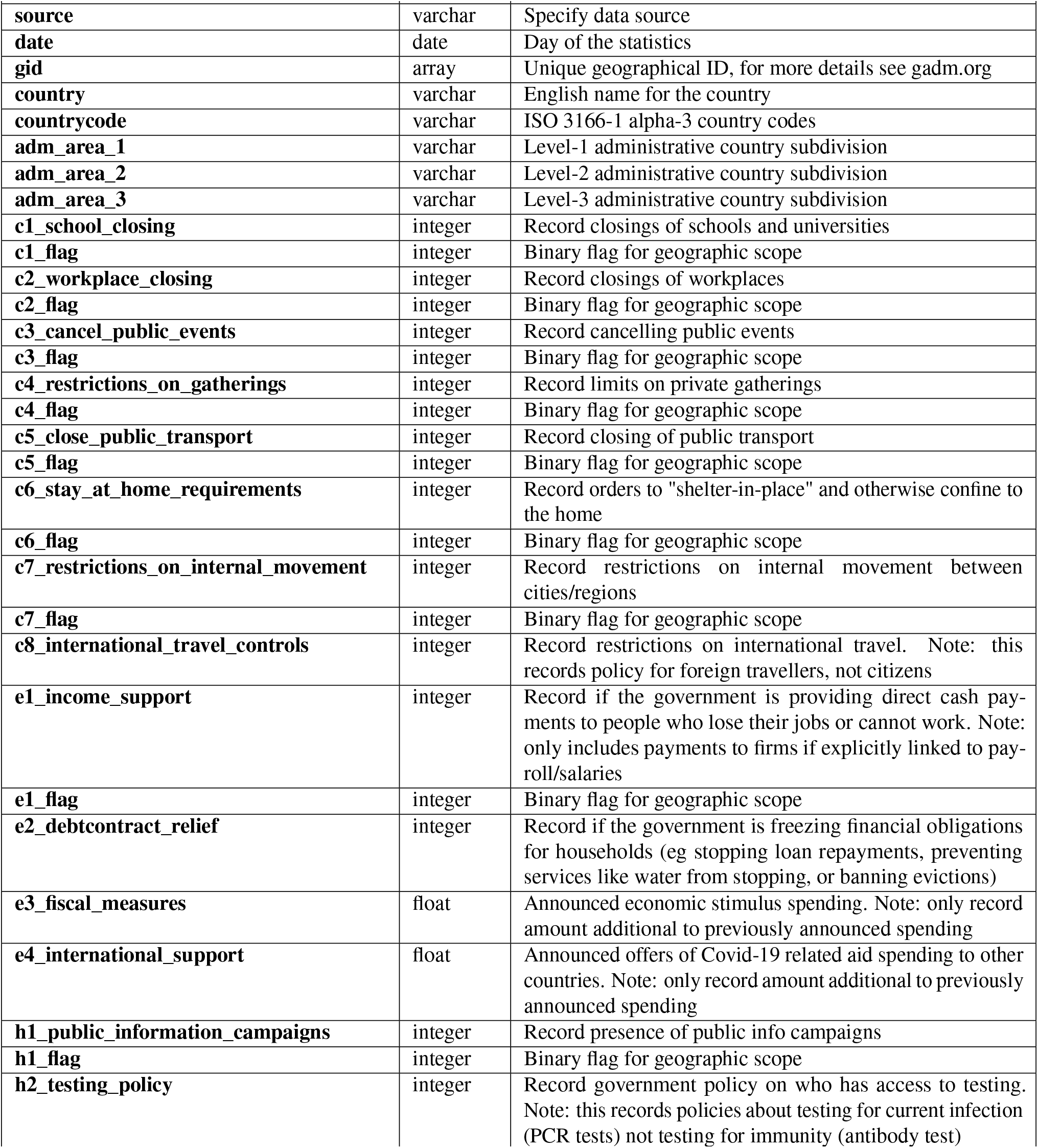
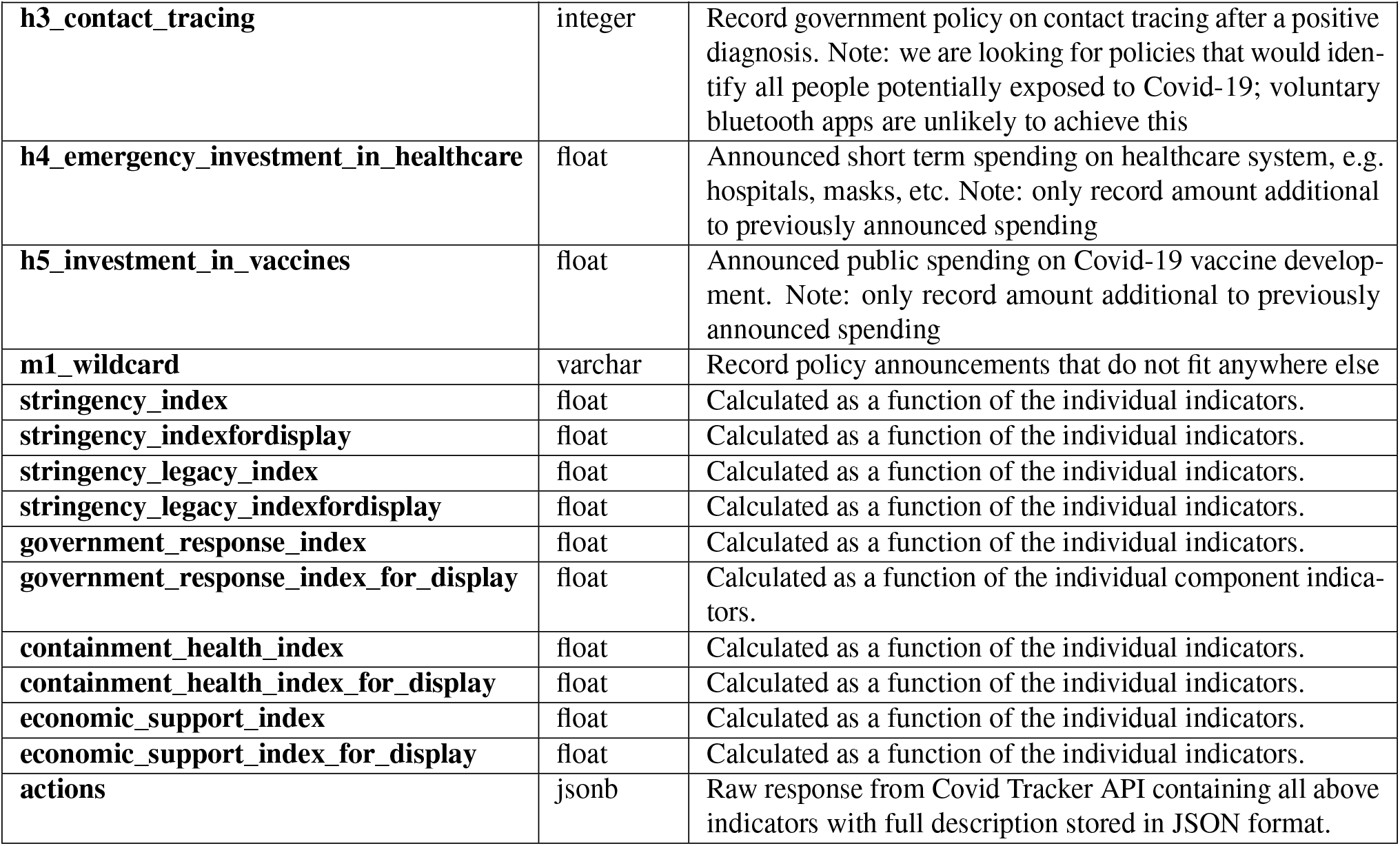
Schema for GOVERNMENT_RESPONSE table.

### Mobility data

The MOBILITY table (see Table 4) includes all eight linking columns, (K1)-(K8), followed by a number of indicators of human mobility as reported by Google^8^. These data are derived from aggregated movements of Android phone users and are stratified by the location of the user: place of work, outdoor parks, recreation areas, grocery markets etc. This table also contains the change in traffic volume reported by Apple^9^ from aggregated tracking of iPhone users of people walking, driving or taking public transit in their communities.

**Table 4.** Schema for MOBILITY table.

| Name | Type | Short description |
| --- | --- | --- |
| **source** | varchar | Specify data source |
| **date** | date | Day of the statistics |
| **gid** | array | Unique geographical ID, for more details see gadm.org |
| **country** | varchar | English name for the country |
| **countrycode** | varchar | ISO 3166-1 alpha-3 country codes |
| **adm_area_1** | varchar | Level-1 administrative country subdivision |
| **adm_area_2** | varchar | Level-2 administrative country subdivision |
| **adm_area_3** | varchar | Level-3 administrative country subdivision |
| **transit_stations** | float | Mobility trends reported by Google for transit stations |
| **residential** | float | Mobility trends reported by Google for places of residence |
| **workspace** | float | Mobility trends reported by Google for places of work |
| **parks** | float | Mobility trends reported by Google for places like parks, national parks, public beaches, marinas, dog parks, plazas and public gardens |
| **retail_recreation** | float | Mobility trends reported Google for places like restaurants, cafes, shopping centers, theme parks, museums, libraries, and movie theaters |
| **grocery_pharmacy** | float | Mobility trends reported by Google for places like grocery markets, food warehouses, farmers markets, specialty food shops, drug stores, and pharmacies |
| **transit** | float | The change in volume reported by Apple of people taking public transit in their communities |
| **walking** | float | The change in volume reported by Apple of people walking in their communities |
| **driving** | float | The change in volume reported by Apple of people driving taking public transit in their communities |

Google measures mobility on any day relative to the median value for each of the five days falling on the same day of the week in the period January 3 – February 6, 2020, while Apple measures all data relative to January 13, 2020. The data only describe mobility within particular locations for particular activities. They do not indicate the amount of travel between regions nor do they contain individual-level data.

### Weather data

The WEATHER table (See Table 5) includes all eight linking columns (K1)-(K8) followed by 47 variables including temperature, sunshine, precipitation, air temperature, wind speed etc.

**Table 5.** Schema for WEATHER table.

| Name | Type | Description |
| --- | --- | --- |
| source | varchar | Specify data source |
| date | date | Day of the statistics |
| gid | array | Unique geographical ID, for more details see gadm.org |
| country | varchar | English name for the country |
| countrycode | varchar | ISO 3166-1 alpha-3 country codes |
| adm_area_1 | varchar | Level-1 administrative country subdivision |
| adm_area_2 | varchar | Level-2 administrative country subdivision |
| adm_area_3 | varchar | Level-3 administrative country subdivision |
| samplesize | int | Number of grid points |
| precipitation_max_avg | float | Average of the daily maximum precipitation |
| precipitation_max_std | float | Standard deviation of the daily maximum precipitation |
| precipitation_mean_avg | float | Average of the daily mean precipitation |
| precipitation_mean_std | float | Standard deviation of the daily mean precipitation |
| humidity_max_avg | float | Average of the daily maximum specific humidity |
| humidity_max_std | float | Standard deviation of the daily maximum specific humidity |
| humidity_mean_avg | float | Average of the daily mean specific humidity |
| humidity_mean_std | float | Standard deviation of the daily mean specific humidity |
| humidity_min_avg | float | Average of the daily minimum specific humidity |
| humidity_min_std | float | Standard deviation of the daily minimum specific humidity |
| sunshine_max_avg | float | Average of the daily maximum short wave radiation |
| sunshine_max_std | float | Standard deviation of the daily maximum short wave radiation |
| sunshine_mean_avg | float | Average of the daily minimum short wave radiation |
| sunshine_mean_std | float | Standard deviation of the daily minimum short wave radiation |
| temperature_max_avg | float | Average of the daily maximum temperature |
| temperature_max_std | float | Standard deviation of the daily maximum temperature |
| temperature_mean_avg | float | Average of the daily mean temperature |
| temperature_mean_std | float | Standard deviation of the daily mean temperature |
| temperature_min_avg | float | Average of the daily minimum temperature |
| temperature_min_std | float | Standard deviation of the daily minimum temperature |
| windgust_max_avg | float | Average of the daily maximum wind gust |
| windgust_max_std | float | Standard deviation of the daily maximum wind gust |
| windgust_mean_avg | float | Average of the daily mean wind gust |
| windgust_mean_std | float | Standard deviation of the daily mean wind gust |
| windgust_min_avg | float | Average of the daily minimum wind gust |
| windgust_min_std | float | Standard deviation of the daily minimum wind gust |
| windspeed_max_avg | float | Average of the daily maximum wind speed |
| windspeed_max_std | float | Standard deviation of the daily maximum wind speed |
| windspeed_mean_avg | float | Average of the daily mean wind speed |
| windspeed_mean_std | float | Standard deviation of the daily mean wind speed |
| windspeed_min_avg | float | Average of the daily minimum wind speed |
| windspeed_min_std | float | Standard deviation of the daily minimum wind speed |
| cloudaltitude_max_valid | float | Percentage of points with a valid value of cloudaltitude_max |
| cloudaltitude_max_avg | float | Average of the daily maximum cloud base altitude |
| cloudaltitude_max_std | float | Standard deviation of the daily maximum cloud base altitude |
| cloudaltitude_min_valid | float | Percentage of points with a valid value of cloudaltitude_min |
| cloudaltitude_min_avg | float | Average of the daily minimum cloud base altitude |
| cloudaltitude_min_std | float | Standard deviation of the daily minimum cloud base altitude |
| cloudaltitude_mean_valid | float | Percentage of points with a valid value of cloudaltitude_mean |
| cloudaltitude_mean_avg | float | Average of the daily mean cloud base altitude |
| cloudaltitude_mean_std | float | Standard deviation of the daily mean cloud base altitude |
| cloudfraction_max_avg | float | Average of the daily maximum cloud area fraction |
| cloudfraction_max_std | float | Standard deviation of the daily maximum cloud area fraction |
| cloudfraction_min_avg | float | Average of the daily minimum cloud area fraction |
| cloudfraction_min_std | float | Standard deviation of the daily minimum cloud area fraction |
| cloudfraction_mean_avg | float | Average of the daily mean cloud area fraction |
| cloudfraction_mean_std | float | Standard deviation of the daily mean cloud area fraction |

**Table 6.**
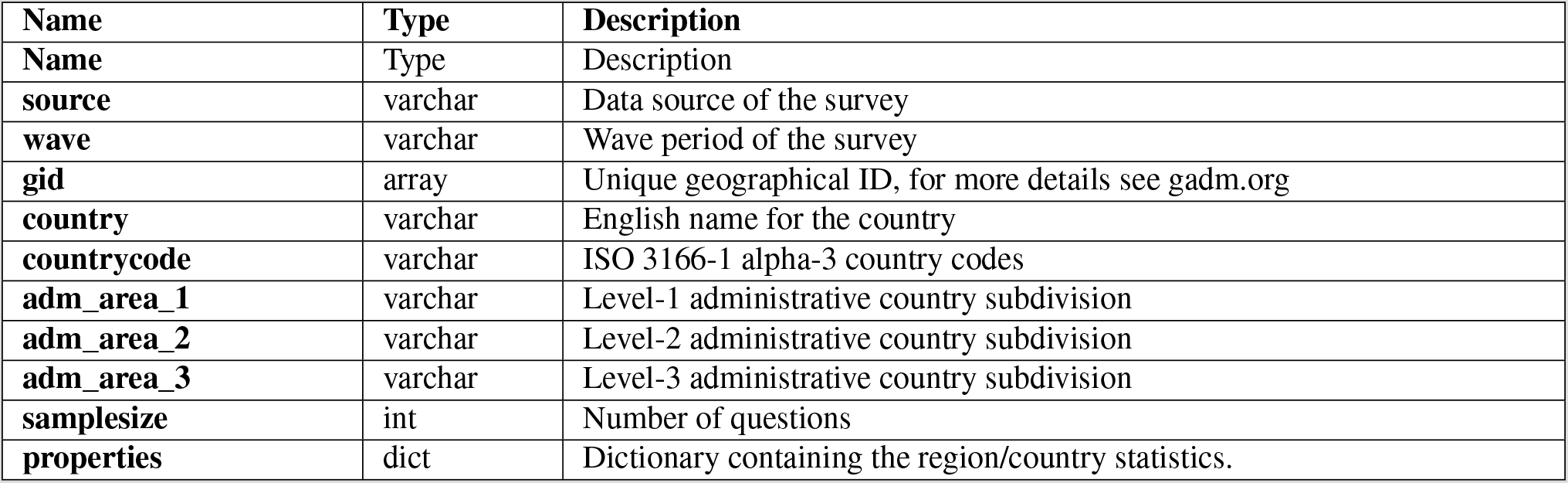
Schema for SURVEYS table.

### World Bank

The WORLD_BANK table (see Table 7) includes seven of the eight linking columns (K1), (K3)-(K8) followed by the **indicator_name, indicator_code, value** and **year**. Each indicator name and the corresponding code relate to one of the 1431 features listed at https://data.worldbank.org/indicator/. The original source provides a series of values from 1960 to 2019. However, here we report only the most recent available value with its year.

**Table 7.** Schema for WORLD_BANK table.

| **Name** | **Type** | **Description** |
| --- | --- | --- |
| **source** | varchar | Specify data source |
| **gid** | array | Unique geographical ID, for more details see gadm.org |
| **country** | varchar | English name for the country |
| **countrycode** | varchar | ISO 3166-1 alpha-3 country codes |
| **adm_area_1** | varchar | Level-1 administrative country subdivision |
| **adm_area_2** | varchar | Level-2 administrative country subdivision |
| **adm_area_3** | varchar | Level-3 administrative country subdivision |
| **indicator_name** | varchar | Description of the indicator |
| **indicator_code** | varchar | World Bank indicator code |
| **value** | float | Most recent non-empty value |
| **year** | int | Year of the most recent value |

### Surveys

The SURVEYS table (see Table 6) includes seven of the eight linking columns (K1), (K3)-(K8) followed by **samplesize** indicating the number of people taking part in the survey for the region under consideration, **properties**, which is a dictionary containing the region/country statistics and **wave**, specifying the particular survey being reported.

## Technical Validation

The code used to build the OxCOVID19 Database was developed collaboratively. Working across several GitHub repositories (https://github.com/covid19db) allowed us to share documentation and keep code organised and up to date. We encourage the research community to report any issues they find. Figure 3 shows the system architecture that is being used to collect, unify, store and share the data. We operate more than 70 fetchers to periodically obtain raw data from our sources. This automated process ensures that we collect the most recent data and reduces potential error due to manual entry. The “Unification” step ensures that the names in different tables in the OxCOVID19 Database are consistent across geographical regions. In the “Validation” step a check for consistency is performed. During the storing step, the last timestamp in input data is compared with the current time and if the inserted data are older than 14 days relevant warnings are generated as that may indicate the change in format of the fetched data or some other problem that needs to be fixed. The fetching process is triggered twice a day at 02:00 and 14:00 BST. The sharing process, namely publishing existing data sources to CSV files hosted on GitHub, is triggered four times a day.

**Figure 3.**
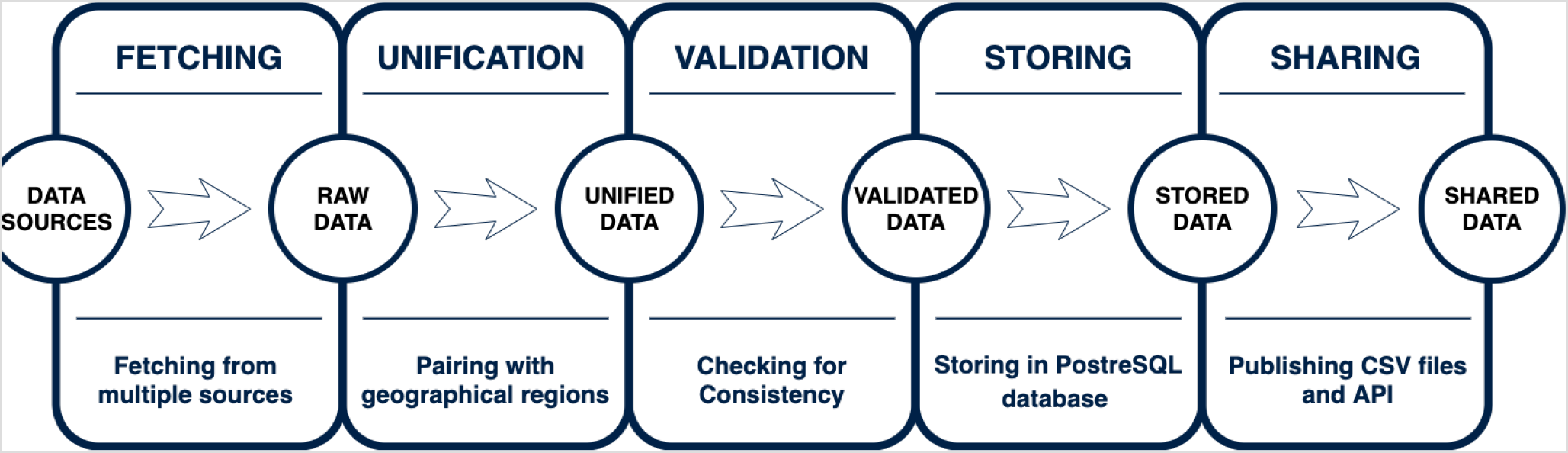
System architecture for OxCOVID19 Database.

## Usage Notes

### Data access

We provide several different means of accessing the OxCOVID19 Database. The latest version can be downloaded in CSV format from https://github.com/covid19db/data or https://covid19.eng.ox.ac.uk/ and the archived static version can be accessed from FigShare https://doi.org/10.6084/m9.figshare.12746150. Direct connection to the PostgreSQL database can also be granted upon request.

### Example usage

The basic examples in Python and R showing how to load the data and perform simple analysis are available at https://github.com/covid19db/examples. We would like to acknowledge that the R package for accessing our database, available on CRAN [https://cran.r-project.org/package=oxcovid19], has been developed by the members of the CoMo Consortium^15^.

### Citation advice

The OxCOVID19 Database is the results of many hours of volunteer efforts and generous contributions from many organisations listed in the Methods section under “Data Sources”. We encourage the users of OxCOVID19 Database to cite, along with this article, the underlying sources.

## Data Availability

The data are stored in a PostgreSQL database. CSV extracts from this database are available to access at https://github.com/covid19db/data. A complete archive copy of the database in CSV format as of 31-07-2020 has been stored under the https://doi.org/10.6084/m9.figshare.12746150.

## Code availability

The code for data acquisition and cleaning used in the processing of assembling the OxCOVID19 Database is on the GitHub repository: https://github.com/covid19db.

## Acknowledgements

We acknowledge the contribution of a number of volunteers and people offering valuable feedback. In particular, we acknowledge the contributions of Abhishek Agarwal, Mario Rubio Chavarría and Tarun Srivastava.

- A.M. and L.T. are funded/supported by the National Institute for Health Research (NIHR) Oxford Biomedical Research Centre (BRC). The views expressed are those of the author(s) and not necessarily those of the NHS, the NIHR or the Department of Health.
- P.D. is supported by the Dioscuri Centre in Topological Data Analysis project financed under Dioscuri – a programme initiated by the Max Planck Society, jointly managed with the National Science Centre in Poland, and mutually funded by Polish Ministry of Science and Higher Education and German Federal Ministry of Education and Research as well as the EPSRC grant New Approaches to Data Science: Application Driven Topological Data Analysis EP/R018472/1.
- N.H. and T.-S.C. is supported by the EPSRC grant New Approaches to Data Science: Application Driven Topological Data Analysis EP/R018472/1.
- J.H. is supported by a Daphne Jackson Fellowship, sponsored by the EPSRC and Swansea University.
- Y.W. acknowledges Alan Turing Institute for funding this work through EPSRC grant EP/N510129/1 and EPSRC through the project EP/S2026347/1, titled ‘Unparameterised multi-modal data, high order signature, and the mathematics of data science’.
- A.E.Z. is supported by Oxford Martin School, Pandemic Genomics programme.
- D.S. is partially funded by the Swedish Knowledge Foundation through the Internet of Things and People research profile.
- B.H. is supported by the US National Institute of Health (R01 DA042711).

## Author contributions statement

A.M. conceived and led the project. P.B. led the technical aspect of the project and developed the solution architecture. A.M., P.B., and P.D. provided the conceptual framework and supervised the development of the database. T.-S.C. and J.H. oversaw the implementation and curation of data fetchers. A.M., P.B., D.S., T.-S.C., J.H., D.G., Y.W., A.F., N.H., and A.Z. wrote data fetchers and examples of use. The individual contributions can be found on GitHub. All authors contributed to the writing of the manuscript.

## Competing interests

The authors declare no competing interests.

## Notes

### Competing Interest Statement

The authors have declared no competing interest.

